# Seroprevalence, correlates and kinetics of SARS-CoV-2 Nucleocapsid IgG antibody in healthcare workers at a tertiary hospital: A prevaccine census study

**DOI:** 10.1101/2022.04.13.22273817

**Authors:** Daniel Maina, Geoffrey Omuse, George Ong’ete, Patrick Mugaine, Shahin Sayed, Zahir Moloo, Reena Shah, Anthony Etyang, Rodney Adam

## Abstract

**Background:** Healthcare workers are perceived to be a high-risk group for acquiring SAR-CoV-2 infection, and more so in countries where COVID-19 vaccination uptake is low. Serosurveillance may best determine the true extent of SARS-CoV-2 infection since most infected HCWs may be asymptomatic or present with only mild symptoms. Over time, determining the true extent of SARS-CoV-2 infection could inform hospital management and staff whether the preventive measures instituted are effective and valuable in developing targeted solutions.

**Methods:** This was a census survey study conducted at the Aga Khan University Hospital, Nairobi, between November 2020 and February 2021 before the implementation of the COVID-19 vaccination. The SARS-CoV-2 nucleocapsid IgG test was performed using a chemiluminescent assay.

**Results:** One thousand six hundred thirty-one (1631) staff enrolled, totalling 60% of the workforce. The overall crude seroprevalence was 18.4% and the adjusted value (for assay sensitivity of 86%) was 21.4% (95% CI; 19.2-23.7). The HCW groups with higher prevalence included pharmacy (25.6%), outreach (24%), hospital-based nursing (22.2%) and catering staff (22.6%). Independent predictors of a positive IgG result included prior COVID-19 like symptoms, odds ratio (OR) 1.9 [95% confidence interval (CI) 1.3-2.9, p=0.002], and a prior positive SARS-CoV-2 PCR result OR 11.0 (CI: 7.2-18.0, p<0.001). Age, sex, comorbidities or working in a COVID-19 designated area were not associated with seropositivity. The odds of testing positive for IgG after a positive PCR test were lowest if the antibody test was performed more than 2 months later; OR 0.7 (CI: 0.48-0.95, p= 0.025).

**Conclusions:** The prevalence of anti-SARS-CoV-2 nucleocapsid IgG among HCWs was lower than in the general population. Staff working in clinical areas were not at increased risk when compared to staff working in non-clinical areas.

## Introduction

SARS-CoV-2 infection remains a threat to public health, especially in low resource settings where vaccine coverage remains low. As of 2^nd^ March 2022, only 13.9% of the Kenyan population was fully vaccinated (1). The positivity rate with PCR testing in the general population continues to fluctuate and was less than 1% at the start of March 2022. Admissions to hospitals reflect this fluctuation. Data on the infection rates of healthcare staff in Kenya are scanty and often do not include details of exposure risk.

The country had 7,466 infected healthcare workers (HCWs) reported as of September 2021. Etyang et al. reported seroprevalences of 43.8% (urban), 12.6% (rural) and 11.5% (rural) in three counties in Kenya (2). The challenges facing HCW on the continent include inadequate personal protective equipment (PPE) and limited SARS-CoV-2 testing of populations that seek medical care, which leaves workers vulnerable (3), as was especially true before the provision of SARS-CoV-2 vaccination to HCW.

Although infections in HCWs are often attributed to occupational exposure, that is not always the case. At Aga Khan University Hospital Nairobi (AKUHN), personal protective equipment (PPE) appropriate for the level of clinical care has been routinely provided since the beginning of the Kenyan outbreak. In addition, routine tests of admitted patients were implemented. Therefore, we wished to know the level of risk to HCWs where PPE and testing are readily available.

Although liberal PCR testing of HCWs has been done at AKUHN throughout the outbreak, asymptomatic HCWs were not routinely tested. Since asymptomatic infections have comprised a significant percentage of infections in some series, it is possible that a significant number of staff infections have been missed (4). The serosurvey helped address the suitability of our approaches to staff safety.

## Materials and Methods

This research was a census study in which all HCW at AKUHN were eligible to participate. The staff were sensitised about the study through posters, institutional email addresses and group talks. One thousand, six hundred thirty-one staff consented and were recruited in the study (>60% of the workforce). The study lasted from November 2020 to February 2021, before the implementation of COVID-19 vaccination in the hospital.

Staff were categorised into five groups based on the perceived risk of COVID-19 exposure at the workplace

✤ Clinical COVID-19 areas: COVID-19 isolation wards, Intensive Care Unit (ICU), High Dependence Unit (HDU), Accident and Emergency (A&E) triage
✤ Non-COVID-19 clinical areas: general wards, outpatient clinics
✤ Allied health: laboratory, radiology, pharmacy
✤ Support staff: catering, facilities, housekeeping
✤ Academic/Administration areas

### Specimen collection & assay

A phlebotomist collected 5-10 ml of blood in a serum separator vacutainer (Greiner Bio-One GmbH – Germany) from each participant. In all cases, blood samples were labelled with a unique study identifier, centrifuged and serum separated within 24 hours of collection.

### Laboratory testing

The antibody test was performed using a chemiluminescent assay (Abbott, USA), according to the manufacturer’s instructions and standard operating procedures. The Abbott qualitative anti-nucleocapsid CoV-2 IgG assay for SARS has a manufacturer’s stated sensitivity of 100% 14 days after the onset of symptoms and a specificity of 99.6%. Our laboratory evaluation using 54 samples from known COVID-19 patients showed a sensitivity of 86%, 14 days after the onset of symptoms, and a specificity of 100% using 20 samples before COVID-19 (unpublished data). An independent evaluation of this kit by Public Health England reported a sensitivity of 93.9% for samples ≥ 14 days post-symptom onset of symptoms and a specificity of 100% (5).

The default result unit for the SARS-CoV-2 IgG assay is Index (S/C), with a cutoff of 1.40. This study was conducted according to the criteria set by the declaration of Helsinki on Human research. The protocol was approved by the Ethics Review Committee of Aga Khan University Nairobi (Ref: 2020/IERC-129 v4).

Written informed consent was obtained from all participants.

### Data collection and Analysis

Stata 16 (StataCorp, Texas, USA) was used to perform the statistical analysis. The demographic and risk details of the patients were captured by filling in a guided questionnaire. Descriptive statistics are presented as medians [interquartile range (IQR)] or means (standard deviation, SD) and percentages (proportions), where appropriate. Crude seroprevalence figures were adjusted for the performance (sensitivity 86%, specificity 100%) of the nucleocapsid assay in our evaluation (6),.

Risks (associations) were analysed using logistic regression analyses and chi-square statistics, presented here, respectively, as odds ratio (OR) with 95% confidence interval (95% CI) and chi statistics with p-value.

Antibody kinetics were evaluated for participants with documented SARS-CoV-2 PCR results and compared with the proportions of HCWs with positive IgG at different time points between the PCR and antibody tests.

## Results

One thousand six hundred thirty-one (1,631) staff members, both AKUHN and contracted employees, participated in the study. This figure represented 60% of the hospital workforce. The median age was 35 years (interquartile range 30-45). AKUHN employees comprised 86.7% of the study participants. Males made up 44% of the tested staff, and the overall crude seroprevalence of SARS-CoV-2 in the study was 18.4%, adjusted to 21.4% (95% CI; 19.2-23.7).

The seroprevalence among AKUHN staff (22.0%) was not significantly different from that of contracted workers (17.7%) (p-value = 0.193). The proportion of staff with SARS-CoV-2 antibodies varied between the different cadres within these two broad groups. Notable cadres with higher proportions of exposed personnel included pharmacy (25.6%), outreach (24%), nursing (22.2%) and catering (22.6%), as shown in Table 1.

**Table 1:** SARS-CoV-2 anti-nucleocapsid IgG antibody prevalence in HCWs.

|  | Total | Seropositive<br>n (crude prevalence) | Adjusted Prevalence<br>(95% confidence<br>interval) |
| --- | --- | --- | --- |
| **All** | 1631 | 300 (18.4%) | 21.4 (19.2-23.7) |
| **Males** | 716 | 126 (17.6%) | 20.5 (17.4-24.0) |
| **Females** | 915 | 174 (19.0%) | 22.1 (19.3-25.2) |
| **Age** |  |  |  |
| 20-29 | 368 | 72 (19.6%) | 22.8% (18.4-27.8) |
| 30-39 | 666 | 124 (18.6%) | 21.6% (18.4-26.2) |
| 40-49 | 392 | 72 (18.4%) | 21.4% (17.2-25.0) |
| 50-59 | 145 | 27 (18.6%) | 21.6% (15.2-30.0) |
| ≥60 | 45 | 3 (6.7%) | 7.8% (2.5-21.8) |
| **Symptomatic**<br>**(preceding 3**<br>**months)** |  |  |  |
| Yes | 750 | 189 (25.2%) | 29.3% (26.1-32.7) |
| No | 881 | 111 (12.6%) | 14.7% (12.4-17.2) |
| **AKU Staff** | 1414 | 267 (18.9%) | 22.0% (19.8-24.4) |
| **Contracted Staff** | 217 | 33 (15.2%) | 17.7% (12.8-24.0) |
| **AKUHN Divisions** |  |  |  |
| Nursing | 347 | 77 (22.2%) | 25.8 (21.0-31.3) |
| Pharmacy | 43 | 11 (25.6%) | 29.8 (14.8-40.6) |
| Outreach | 117 | 28 (24.0%) | 27.9 (19.9-37.8) |
| Pathology | 84 | 14 (16.7%) | 19.4 (11.7-30.5) |
| Diagnostic imaging | 53 | 9 (17.0%) | 19.8 (10.6-34.3) |
| Medical services | 466 | 78 (16.7%) | 19.4 (15.7-23.7) |
| Others (nonclinical) | 304 | 57 (18.8%) | 21.9 (16.9-27.4) |
| **Risk Based** |  |  |  |
| Clinical Covid | 209 | 49 (23.4%) | 27.2 (21.2-34.5) |
| Non-Covid Clinical | 530 | 99 (18.7%) | 21.7 (18.1-25.9) |
| Allied health | 234 | 37 (15.8%) | 18.4 (13.6-24.5) |
| Support | 185 | 35 (18.9%) | 22.0 (16.2-29.3) |
| Admin/Academic | 249 | 46 (18.5%) | 21.5 (16.4-27.7) |
| **Contracted staff** |  |  |  |
| Catering | 53 | 12 (22.6%) | 26.3 (14.3-42.1) |
| Housekeeping | 103 | 14 (13.6%) | 15.8 (8.8-25.3) |
| Security | 35 | 4 (11.4%) | 13.3 (3.7-31.0) |
| Others | 26 | 3 (11.5%) | 13.4 (2.9-35.1) |

**Key:** Clinical Covid: isolation wards, ICU, HDU, A&E triage; Clinical non-Covid: general wards, outpatient clinics; Allied health: laboratory, radiology, pharmacy; Support staff: catering, facilities, housekeeping; Academic/Administration: nonclinical faculty, administrative staff

Seven hundred sixty-nine HCW (47%) had taken a SARS-CoV-2 PCR test prior to the study, with 136 (18%) confirmed positive. Seroprevalence was 66.2% among HCWs with a positive PCR test and 13.5 % among those with a negative result (p<0.001). Slightly less than half (46%) of the 1631 HCW reported symptoms consistent with COVID-19 in the previous 3 months prior to the study. The most prevalent symptoms were headache (52%), sneezing (51%), cough (42%) and fever (16%). Adjusted seroprevalence among symptomatic and asymptomatic HCWs was 29.3% and 14.7%, respectively (p<0.001).

### Predictors of a positive anti-SARS-CoV-2 nucleocapsid IgG test

Having previously tested for COVID-19 (PCR) irrespective of test result increased the odds of testing positive with the antibody test, OR 1.7 (95% CI: 1.3-2.2, p <0.001) (Table 2). The odds were even higher in those who had tested positive with PCR (OR 12.5 95% CI: 8.2-19.1, p<0.001). This also applied to those who had experienced flu-like symptoms in the previous 3 months prior to enrollment in the study, OR 2.3 (95% CI 1.8-3.0, p<0.001); antibody test performed 2-4 weeks after the PCR test, OR 1.6 (95% CI 1.02-2.54, p =0.041); or reported daily contact with COVID-19 patients, OR 1.5 (95% CI 1.2-2.0, p=0.002).

**Table 2:** Risk factors for a positive anti-nucleocapsid IgG result.

|  | SARS-CoV-2 IgG<br>Positive<br>n (%) | Odds Ratio (95%<br>CI) | p-value |
| --- | --- | --- | --- |
| **Previously tested**<br>**for SARS-CoV-2**<br>**(PCR)** |  |  |  |
| Yes | 175 (22.8) | 1.7 (1.3-2.2) | <0.001 |
| No | 124 (14.5) |  |  |
| **Time interval**<br>**between PCR and**<br>**serology test** |  |  |  |
| <2 weeks | 25 (23.6) | 1.1 (0.6-1.7) | 0.855 |
| 2-4 weeks | 32 (30.8) | 1.6 (1.02-2.54) | 0.041 |
| 1-2 months | 37 (25.7) | 1.2 (0.8-1.8) | 0.375 |
| >2 months | 80 (19.7) | 0.7 (0.48-0.95) | 0.025 |
| **COVID-19 PCR**<br>**result** |  |  |  |
| Positive | 90 (66.2) | 12.5 (8.2-19.1) | <0.001 |
| Negative | 85 (13.5) |  |  |
| **Flu-like symptoms**<br>**past 3 months** |  |  |  |
| Yes | 189 (25.2) | 2.3 (1.8-3.0) | <0.001 |
| No | 111 (12.6) |  |  |
| **Univariate analysis**<br>**of individual**<br>**symptoms** |  |  |  |
| Cough | 93 (29.6) | 2.3 (1.7-3.0) | <0.001 |
| Sneezing | 92 (24.2) | 1.6 (1.2-2.1) | 0.001 |
| Fever | 50 (42.4) | 3.7 (2.5-5.5) | <0.001 |
| Running nose | 87 (24) | 1.6 (1.2-2.1) | 0.002 |
| Chest pain | 29 (26.6) | 1.7 (1.1-2.6) | 0.023 |
| Headache | 102 (26) | 1.8 (1.4-2.4) | <0.001 |
| Loss of smell | 59 (67.8) | 11.4 (7.1-18.2) | <0.001 |
| **Multivariate**<br>**analysis symptoms** |  | Adjusted OR |  |
| Cough |  | 1.7 (1.2-2.4) | 0.004 |
| Fever |  | 1.7 (1.1-2.8) | 0.026 |
| Loss of smell |  | 8.6 (5.2-14.3) | <0.001 |
| **Household contacts or workmates diagnosed with covid** |  |  |  |
| Yes | 163 (19.6) | 1.2 (0.9-1.5) | 0.220 |
| No | 137 (17.2) |  |  |
| **Work involves frequent (daily) close contact with COVID-19 patients** |  |  |  |
| Yes | 190 (21.1) | 1.5 (1.2-2.0) | 0.002 |
| No | 110 (15.1) |  |  |
| **Hospital Location Risk (clinical COVID)** |  |  |  |
| Yes | 49 (22) | 1.3 (0.9-1.8) | 0.171 |
| No | 251 (18) |  |  |

Univariate analysis of specific symptoms showed a significant association between cough, headache, fever, coryza and loss of smell with the positive Covid-IgG test. However, multivariate analysis revealed an independent association with only cough, fever and loss of smell.

Age, sex, and comorbidities (diabetes and hypertension) were not risk factors for a positive SARS-CoV-2 IgG test.

A binomial logistic regression was run to understand the effects of a positive COVID-19 PCR result, flu-like symptoms 3 months prior to testing, working in close contact with COVID-19 patients and antibody test performed 2-4 weeks after PCR test on the likelihood of being seropositive. A positive PCR result and flu-like symptoms predicted seropositivity. However, interacting daily with COVID-19 patients and the time interval between PCR and antibody tests did not achieve statistical significance (Table3).

**Table 3:** Predictors of a positive anti-nucleocapsid IgG result.

|  | Univariate |  | Multivariate Model<sup>a</sup> |  |
| --- | --- | --- | --- | --- |
|  | Odds Ratio<br>(95% CI) | p-value | Odds Ratio<br>(95% CI) | p-value |
| SARS-CoV-2 PCR result + | 12.5 (8.2-19.1) | <0.001 | 11.0 (7.2-17.0) | <0.001 |
| Flu-like symptoms past 3 months | 2.32 (1.8-3.0) | <0.001 | 1.9 (1.3-2.9) | 0.002 |
| Work involves frequent (daily) close contact with COVID-19 patients | 1.5 (1.2-2.0) | 0.002 | 1.3 (0.8-1.8) | 0.267 |
| Time interval (2-4 weeks) Between PCR and antibody test | 1.6 (1.02-2.54) | 0.041 | 1.2 (0.7-2.1) | 0.450 |

<sup>a</sup>Model includes risk factors with p< 0.10 on univariate analysis: positive SARS-CoV-2 PCR result, presence of flu-like symptoms, frequent close contact with COVID-19 patients at the workplace and time between the PCR and antibody test

### COVID-19 IgG dynamics

The seroprevalence in participants who tested positive for SARS-CoV-2 by PCR peaked 1-2 months following the PCR test and sharply declined thereafter (Figs 1&2). The odds of testing antibody positive were lowest if performed more than 2 months after PCR, OR 0.7 (CI 0.48-0.95). The PCR negative group had a steady but slow decline in seroprevalence throughout the same period.

**Fig 1.**
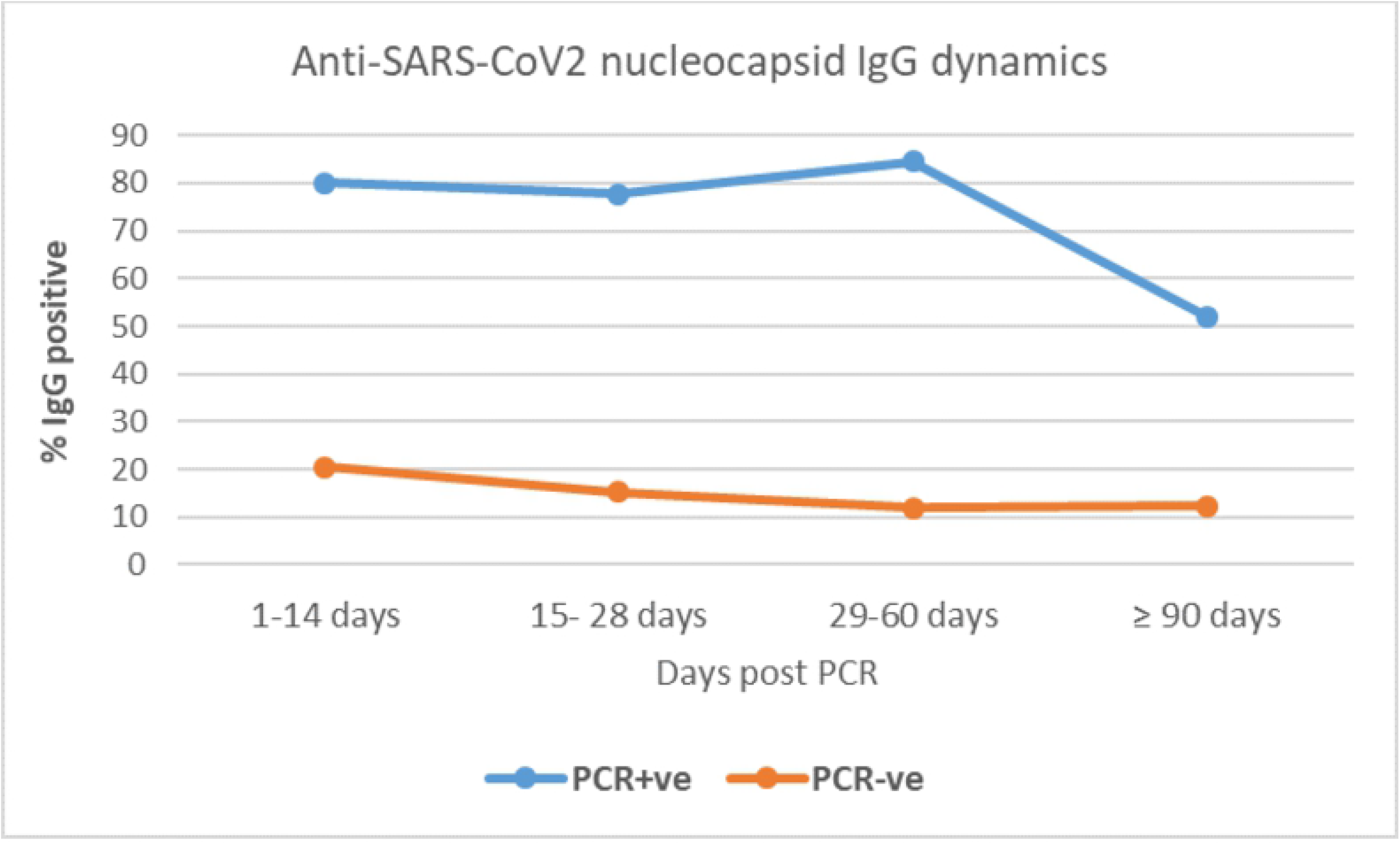
Proportion of HCWs with positive antibody over time after PCR test.

**Fig 2.**
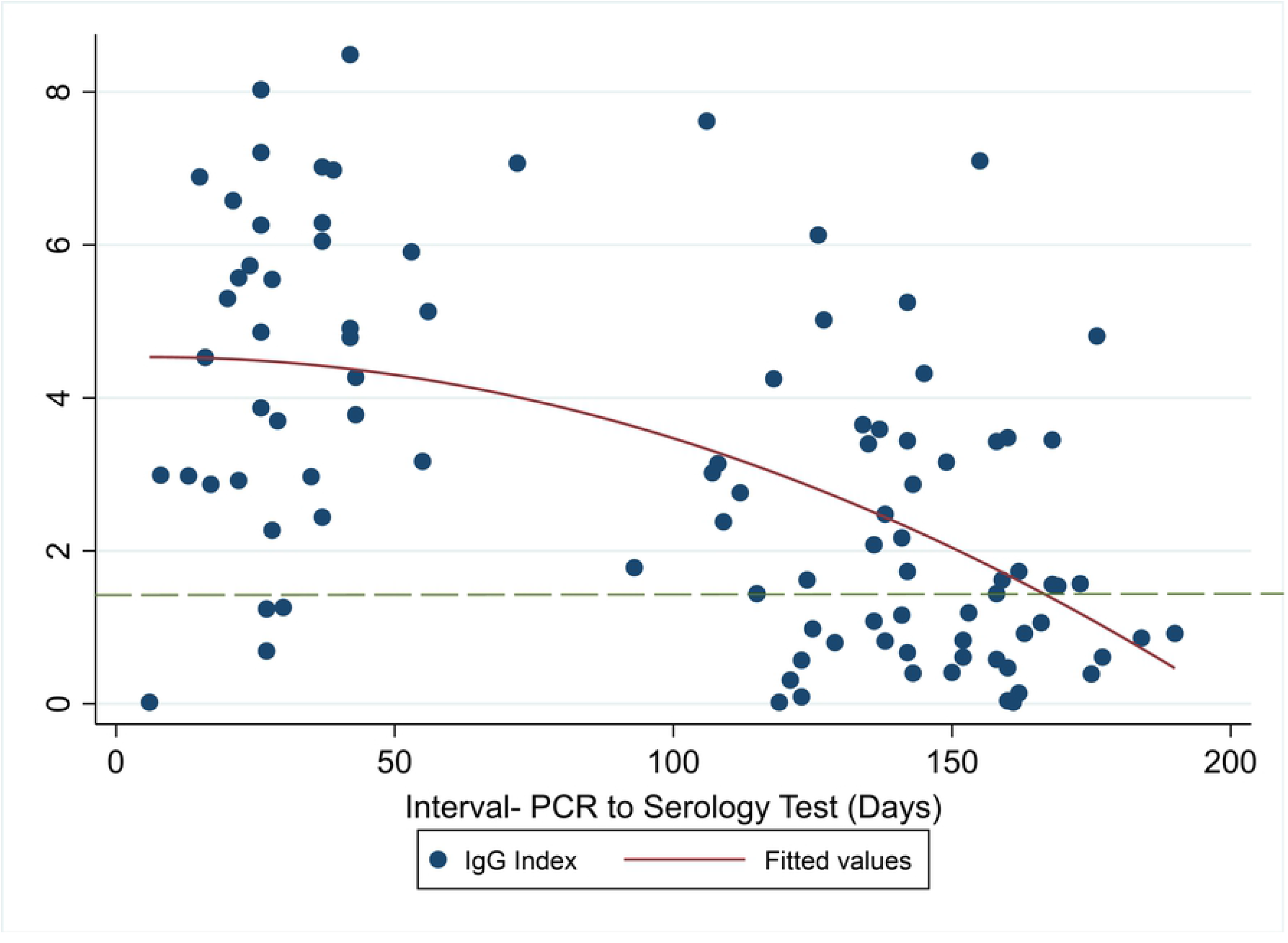
Antibody index at various time points post-SARS-CoV-2-19 PCR+ test. Key: The dashed horizontal line is the assay cutoff point (1.4)

## Discussion

We determined the seroprevalence and kinetics of SARS-CoV2 nucleocapsid IgG antibodies in HCW prior to the advent of the COVID-19 vaccine when little information was available on the clinical implications (immunity) of seroconverting. This was also before any variant of concern had been reported in the country. The adjusted seroprevalence of 21% was well below any reported in a public urban hospital (43.8%) in Nairobi (2). Whether the strict COVID-19 prevention measures enforced early in AKUHN contributed substantially to the relatively lower seroprevalence has not been determined. Hospitals in rural areas had lower rates reflecting low community transmission (2). Early in the COVID-19 pandemic, Nairobi was the epicentre of the infection, which later spread to other parts of the country.

Elsewhere on the continent, seroprevalence among HCW ranged from 0% to 45.1%, with the highest prevalence in Nigeria (7).

Contrary to previous assumptions, there was no significant difference in seroprevalence between frontline staff (high-risk work-based exposure) and staff working in nonclinical areas (low-risk work exposure), suggesting that most of the SARS-CoV-2 infections occurred in the community. During the same period, a study conducted in India reported similar figures between HCWs and the general public (25.6% versus 23.2%) (8). Here in Kenya, a study reported a higher prevalence (61.8%) among blood donors than the prevailing seroprevalence among HCWs in Nairobi for the period January through March 2021(9), reflecting the high community transmission prevalent during the study period.

The seroprevalence across the age groups was similar for HCW under 60 years of age and then decreased for age ≥ 60. Some studies have found increased seropositivity with increasing age for some age brackets (10, 11). Pharmacy and catering staff had the highest prevalence, possibly explained by confined working spaces with close contact between the staff in the two areas.

Testing positive for SARS-CoV-2 by PCR and having flu-like symptoms independently predicted a positive SARS-CoV-2 antibody result. The three symptoms that independently predicted a positive antibody result were loss of smell, followed by fever and cough (adjusted OR 8.7, 1.7, 1.7, respectively). Other studies have reported similar symptoms among HCWs elsewhere (12, 13).

Symptomatic staff had mild COVID-19 disease, none of which required hospitalisation. The hospital workforce is relatively young (mean age of 37 years) and has few comorbidities, which could partly explain the mild COVID-19 disease seen in the facility.

Correct interpretation of serology test results requires a proper understanding of the antibody dynamics, as demonstrated in our findings. Testing for nucleocapsid IgG antibodies three months after SARS-CoV-2 infection is likely to miss a significant proportion of exposed people. Only 20% of HCW with confirmed infection in our study were positive for IgG antibodies at three months and beyond. Van Elslande et al. reported that anti-nucleocapsid IgG antibody levels of SARS-CoV-2 steadily decreased after 2 months up to 8 months after PCR, while another study showed a decline in positivity starting at 6 months onward after the onset of COVID-19 symptoms or testing (14, 15).

All participants in our study had mild disease or were asymptomatic and the kinetics described here may not apply to those with severe COVID-19 disease, which has been associated with earlier and higher antibody levels compared with asymptomatic infections (16, 17).

## Conclusions

The prevalence of anti-SARS-CoV-2 nucleocapsid IgG among HCWs was high but lower than in the general population. There were no statistically significant differences in seroprevalence between the clinical areas and the rest of the hospital. Independent predictors of a seropositive result were a positive SARS-CoV-2 PCR result and flu-like symptoms in the 3 months prior to joining the study. Seropositivity peaked 2 months after a positive PCR test and then declined.

## Data Availability

Data cannot be shared publicly because of confidential reasons as the data include medical information of employees. Data are available from the Aga Khan University Institutional Ethics Committee (contact via) for researchers who meet the criteria for access to confidential data.

## Acknowledgements

Much appreciation to the administration of AKUHN for facilitating the study. Thanks to Cyrus Matheka, David Kawalya and Assumpta Chege for the logistical assistance.

## Notes

### Competing Interest Statement

The authors have declared no competing interest.

### Funding Statement

The authors received no specific funding for this work

### Author Declarations

The research was approved by the Ethics Review Committee of Aga Khan University Nairobi (Ref: 2020/IERC-129 v4).

